# Integrative observational and genetic evidence from the UK Biobank supports carotid intima-media thickness as a window into cardiac remodeling

**DOI:** 10.1101/2025.11.20.25340640

**Authors:** Si Hui Chong, Adama Saccoh, Marion van Vugt, A. Floriaan Schmidt, Nikhil Paliwal

## Abstract

**Objective:** Carotid intima-media thickness (cIMT) is a non-invasive marker of subclinical atherosclerosis and has been linked to future cardiovascular events. However, its relationship with early cardiac structural and functional remodeling and its shared genetic basis with cardiovascular diseases (CVD) remain poorly defined. We investigated the phenotypic and genetic associations between cIMT, aortic and cardiac imaging traits, and CVD in the UK Biobank.

**Methods:** We analyzed 51,818 participants with both carotid ultrasound and cardiac magnetic resonance (CMR) imaging, excluding individuals with prior heart failure (HF), atrial fibrillation (AF), myocardial infarction (MI), or stroke. Multivariable linear and logistic regression models, adjusted for age, sex, and body mass index, assessed associations between cIMT, aortic and cardiac traits, and CVD outcomes. Genetic correlations were estimated using linkage disequilibrium score regression (LDSC) based on GWAS summary statistics. Gene-level and pathway enrichment analyses were performed to identify shared molecular mechanisms linking cIMT with aortic and cardiac phenotypes.

**Results:** Higher cIMT was significantly associated with larger ascending and descending aortic diameters, increased left ventricular (LV) mass and wall thickness, and greater atrial and ventricular volumes, alongside reduced left and right atrial ejection fractions (all p < 0.001). Elevated cIMT was associated with increased AF risk (OR = 1.39, 95% CI: 1.02 to 1.91, P = 0.04 for left cIMT, OR = 1.90, 95% CI: 1.35 to 2.67, P < 0.001 for right cIMT), and left cIMT additionally predicted HF (OR = 2.05, 95% CI: 1.23 to 3.41, P = 0.006), independent of conventional risk factors. Genetically, cIMT showed strong positive correlations with descending aorta, LV wall thickness, and cardiac chamber volumes, as well as with right ventricular end-diastolic volume, LV cardiac output and LV mass. In contrast, cIMT was negatively correlated with left atrial ejection fraction. Seven shared genes (*CDH13, HAND2, ITCH, FBXO32, TBX20, FBN1*, and *CBFA2AT3)* were identified, enriched in pathways related to extracellular matrix organization, vascular development, and cardiac morphogenesis.

**Conclusion:** Elevated cIMT is independently associated with systemic aortic and cardiac remodeling and impaired atrial and ventricular function related to AF and HF. Integrative genetic analyses reveal shared heritable mechanisms linking vascular thickening and myocardial remodeling, highlighting cIMT as a marker of early cardiovascular structural change and a potential bridge between vascular and cardiac disease processes.

**GRAPHICAL ABSTRACT:** 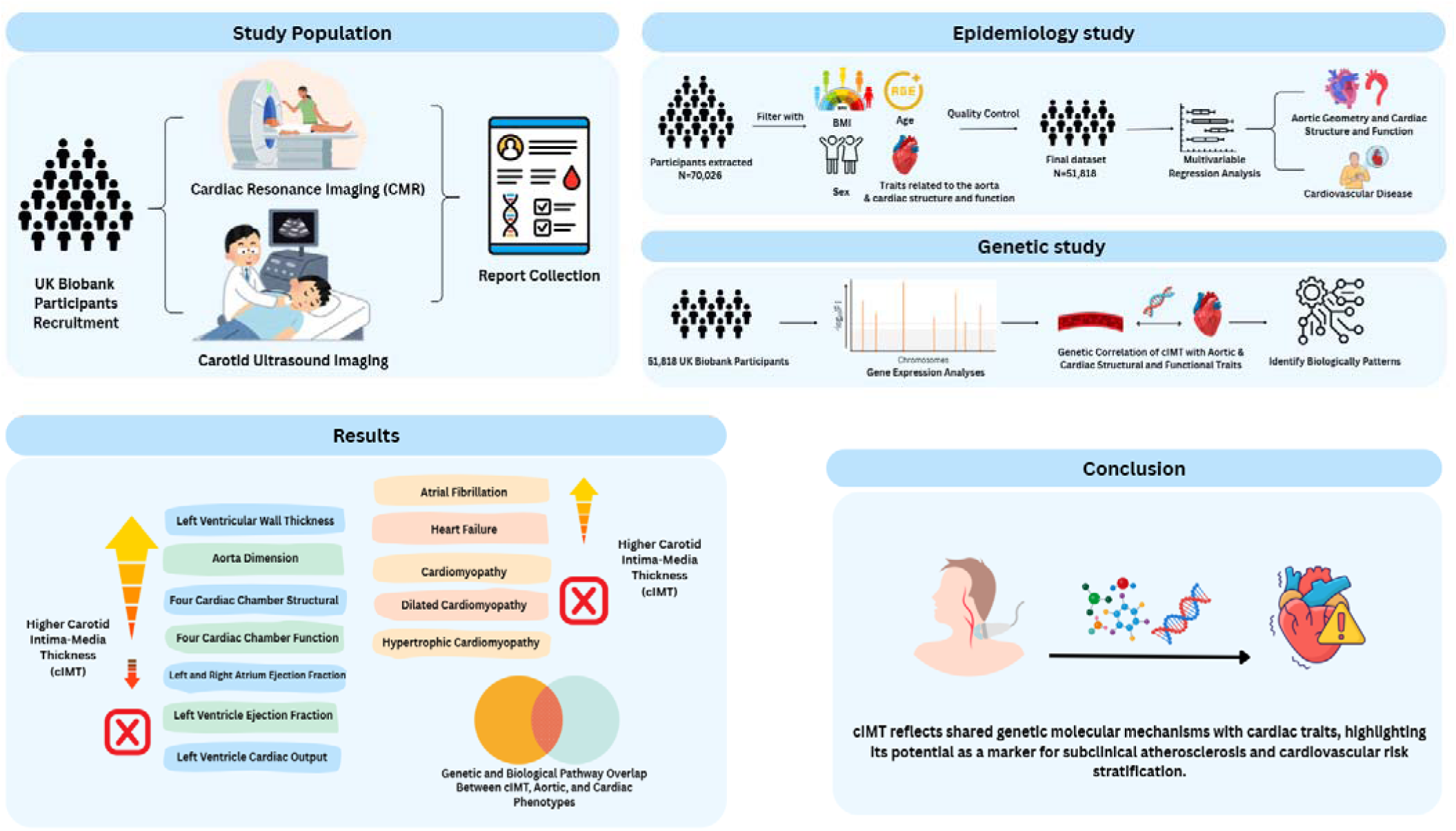

## INTRODUCTION

Atherosclerotic cardiovascular disease (CVD) is a leading global cause of death, with atherosclerosis as the primary underlying pathology.^1^ Atherosclerosis is marked by early lipid accumulation, vascular remodeling, chronic inflammation, and plaque formation. These changes may progress sub-clinically from adolescence and culminate in major cardiovascular events such as MI or stroke.^2,3^ Ultrasound is a widely used non-invasive technique for assessing carotid intima-media thickness (cIMT), a surrogate marker of subclinical atherosclerosis.^4,5^ Large epidemiological cohorts, including the Multiethnic Study of Atherosclerosis (MESA),^6^ the Atherosclerosis Risk in Communities (ARIC) study,^7^ have shown that increased cIMT predicts future cardiovascular events, highlighting its potential utility in early risk stratification.

As a systemic vascular disorder, atherosclerosis also affects major arteries, including the aorta and carotids, with implications for cardiac structure and function.^8^ To examine this, studies have linked cIMT to cardiac morphology and function, as well as to CVD outcomes such as stroke, MI, and coronary heart disease (CHD).^9,10^ However, these investigations have often been limited by small sample sizes, heterogeneous measurement protocols, and incomplete characterization of cardiac and aortic phenotypes.

At the genetic level, genome-wide association studies (GWAS) have identified loci associated with cIMT and related atherosclerotic traits as well as cardiac traits.^11–14^ However, whether cIMT shares a common genetic architecture with cardiac or aortic remodeling traits remains largely unexplored. Understanding such shared heritability could provide insight into the molecular pathways underlying early cardiovascular remodeling and identify potential mechanisms linking subclinical atherosclerosis to overt cardiac disease.

In this study, we performed an integrative epidemiological and genetic analysis using the UK Biobank cohort and publicly available genetic data from the CHARGE (Cohorts for Heart and Aging Research in Genomic Epidemiology).^15^ We first analysed the phenotypic associations between cIMT and aortic and cardiac structure and function, as well as with CVD outcomes in asymptomatic individuals. We explored the shared genetic architecture of cIMT, aortic, and cardiac traits to gather insights into the early molecular and structural pathways linking subclinical atherosclerosis to CVD.

## MATERIALS AND METHODS

### Study sample

We used the UK Biobank, a large-scale prospective cohort study that recruited over half a million individuals aged 40 to 69 years between 2006 and 2010 across the United Kingdom, in this study. Detailed information regarding the UK Biobank cohort has been previously published.^16^ Comprehensive whole-body imaging was available for over 60,000 of these participants.^17^ Cardiac magnetic resonance (CMR) images and B-mode ultrasound were utilised for cardiac characterization and cIMT assessment, respectively.^18,19^ Based on linked clinical records, individuals with a history of MI, angina, heart failure (HF), stroke, cardiomyopathy, or atrial fibrillation (AF), were excluded from the analysis. Individual-level data on age, sex, body mass index (BMI), and plaque status were also extracted from the UK Biobank. Northwest Multi-Center Research Ethics Committee approved ethical approval for the UK Biobank Study. Generic approval for the UK Biobank Study was granted by National Health Service (NHS) National Research Ethics Service (Ref. 11/NW/0382).

### Carotid IMT imaging and extraction

Carotid ultrasound imaging was performed using the CardioHealth Station system (Panasonic Biomedical Sales Europe BV, Leicestershire, UK) with a 9 MHz transducer. Carotid IMT measurements were acquired at fixed angles, 120° and 150° for the right carotid artery, and 210° and 240° for the left, resulting in four cIMT values per participant. cIMT was measured at each acquisition angle, and maximum, minimum, and mean values were recorded at end-diastole.^17^ The average of the four common cIMT measurements was calculated by integrating the maximum values across multiple carotid angles, incorporating data from both the near and far arterial walls. The final mean cIMT value for each participant was calculated as the average across all measured angles, providing a representative estimate of overall carotid wall thickness.^20^ cIMT measurements that did not meet quality control standards, either due to zero values or identified errors, were omitted from the study.^19^

A custom in-house pipeline was developed to extract cIMT data from raw vascular imaging records. An optical character recognition (OCR)-based workflow was implemented to convert image-embedded text into machine-readable outputs.^21^ The script detected predefined text anchors and numerical patterns corresponding to left and right cIMT mean values. To evaluate accuracy, a manual quality control step was performed on a randomly selected subset of 30 imaging records. Detailed information on the extraction workflow is provided in Supplementary Figure S1.

### CMR protocol and image processing

The UK Biobank CMR procedure has been previously described.^22,23,24,25^ For CMR images, we processed the left ventricle (LV) short-axis, long-axis, and aortic images. Each of these images was segmented using a pre-trained neural network to identify the aorta and the 4 cardiac chambers: left atrium (LA), right atrium (RA), LV and right ventricle (RV). Detailed methodology and validation of the segmentation model can be found in Bai et al..^25^ Based on the segmentation, we quantified 25 cardiac traits, spanning the aorta (4 traits), atria (4/4 LA/RA), ventricular (9/4 LV/RV), see Supplementary Table 1 for details.^26^

### Demographic and clinical data

For each participant with cIMT measurement, we extracted key variables, including age, BMI, sex, and CVD outcomes (AF, HF, cardiomyopathy, DCM, and HOCM). For the CVD analyses, participants with a prevalent CVD diagnosis before or within 6 months of the imaging visit were classified as controls to minimise reverse causation and ensure focus on subclinical cardiovascular changes.

### Epidemiology analysis

Characteristics of the participants were explored through stratification by sex. Continuous variables with a normal distribution were summarized as mean (standard deviation, SD), and skewed variables as median (interquartile range, IQR). Categorical variables were presented as percentages (%). Mann-Whitney U test was used to assess significant differences in measurement values between females and males, while the Chi-square test was used to examine the significance of differences in categorical variables between the two groups. Associations between cIMT (independent variable), aortic and cardiac phenotypes, and CVD were examined using multivariable linear and logistic regression models. All primary models were adjusted for age, sex, and BMI.

Separate models were fitted for each dependent variable, including cardiac structural and functional parameters (ventricular volumes, myocardial mass, wall thickness, and atrial volumes and function) and CVD outcomes (AF, HF, cardiomyopathy, DCM, and HOCM). For continuous variables, results are reported as the change in the dependent variable per one standard deviation (SD) increase in the independent variable. For binary variables, results are expressed as the odds of having CVD per one SD increase in the independent variable, with corresponding 95% confidence intervals (CI). The statistical significance threshold was set at *P* < 0.005. Statistical analysis was performed using Python 3.13.2 (https://www.python.org).

### Genetic analysis

#### Genetic correlation between cIMT and cardiac structure and function

Linkage disequilibrium score regression (LDSC, v1.0.1) was used to estimate genetic correlations between cIMT, and a range of aortic and cardiac traits.^27^ GWAS summary statistics for the cIMTmean phenotypes were downloaded from the CHARGE (Cohorts for Heart and Aging Research in Genomic Epidemiology) consortium (http://www.chargeconsortium.com/), and summary statistics for aortic and cardiac traits from the local UCL cohort, respectively.^28^

#### Variant-level annotations and genetic analysis

Genome-wide association results for cIMT-associated, aortic, and cardiac traits were analysed using GWASLab.^29^ Genome-wide significant lead variants, defined as surpassing the significance threshold of P < 5×10⁻ within a 500 kb window, were used to identify a set of independent SNPs associated with cIMT phenotypes, aortic, and cardiac traits. Variants exhibiting –log₁₀(p) values below 2 were omitted to optimize computational efficiency during plotting, while variants with –log₁₀(p) values exceeding 20 were rescaled down to this limit to improve the clarity and interpretability of the visualization. The closest protein-coding gene was annotated for each corresponding lead variant, and the resulting gene set was used to identify genes shared between cIMT, aortic, and cardiac traits.

#### Gene set enrichment and pathway analyses

Functional and pathway enrichment analyses were performed using g:GOSt within the g:Profiler suite, which integrates annotations from Gene Ontology, Human Phenotype Ontology, Reactome, KEGG, and WikiPathways. Genes mapped to validated loci for cIMT, aortic, and cardiac traits were used as input, and results were summarized to highlight enriched biological pathways and molecular processes.^30^

## RESULTS

### Study population

This study included 51,818 participants (51.2% female), with details of the population listed in Table 1. Female participants were slightly younger (mean age 64.4 vs 65.7 years) and had a lower BMI (25.9 vs 26.9 kg/m²). Carotid plaque was more common in men (right 0.05%, left 0.03%) than in women (0.01 each). Males also had a higher prevalence of CVD, including AF (1.9% vs 0.8%), HF (0.6% vs 0.3%), cardiomyopathy (0.12% vs 0.06%), DCM (0.05% vs 0.01%), and HOCM (0.06% vs 0.02%). Mean cIMT values were lower in females than in males, with smaller overall aortic and cardiac dimensions; however, females exhibited slightly higher ejection fraction (p < 0.001). Details of statistical analysis can be found in Supplementary Table 2.

**Table 1.** Characteristics of the UK Biobank participants stratified by sex. Data are reported as mean (SD) for normally distributed and median (Q1, Q3) for non-normally distributed continuous variables; categorial variables are summarised as count (%).

| Variable | Female (N=26,533) | Male (N=25,285) | p-value |
| --- | --- | --- | --- |
| Age (years) | 64.4 (7.6) | 65.7 (7.8) | <0.001 |
| BMI (kg/m<sup>2</sup>) | 25.9 (4.6) | 26.9 (3.8) | <0.001 |
| **cIMT Measurements** |  |  |  |
| Left Mean cIMT | 0.68 (0.60, 0.78) | 0.74 (0.63, 0.87) | <0.001 |
| Right Mean cIMT | 0.68 (0.61, 0.77) | 0.72 (0.62, 0.83) | <0.001 |
| Right Plaque Present | 3 (0.01) | 13 (0.05) | <0.001 |
| Left Plaque Present/Absent | 3 (0.01) | 8 (0.03) | <0.001 |
| **Cardiovascular Disease** |  |  |  |
| Atrial Fibrillation | 404 (0.8) | 957 (1.9) | <0.001 |
| Heart Failure | 128 (0.3) | 330 (0.6) | <0.001 |
| Cardiomyopathy | 32 (0.06) | 56 (0.12) | 0.09 |
| Dilated Cardiomyopathy | 6 (0.01) | 26 (0.05) | 0.02 |
| Hypertrophic Cardiomyopathy | 11 (0.06) | 30 (0.02) | 0.054 |

### Associations between cIMT, aortic, and cardiac traits, and CVD

In fully adjusted multivariable regression models, higher left and right mean cIMT were consistently associated with larger aortic cross-sectional areas, increased chamber size, and greater atrial and ventricular volumes (all P < 0.001; Figure 1). Conversely, cIMT demonstrated significant inverse associations with left and right atrial ejection fractions (P < 0.001), indicating early functional impairment accompanying structural enlargement. While higher cIMT correlated positively with left atrial stroke volume (LASV), an inverse association was observed with right atrial stroke volume (RASV) for left-sided cIMT (P= 0.01), whereas right-sided cIMT showed no association (P = 0.5). Right atrial maximum volume (RAVmax) was significantly associated with right cIMT but not with left cIMT (P = 0.06). No significant relationships were observed between cIMT and left ventricular ejection fraction (LVEF) or cardiac output (CO) (all P > 0.05). Further details can be found in Supplementary Table 3.

**Figure 1.**
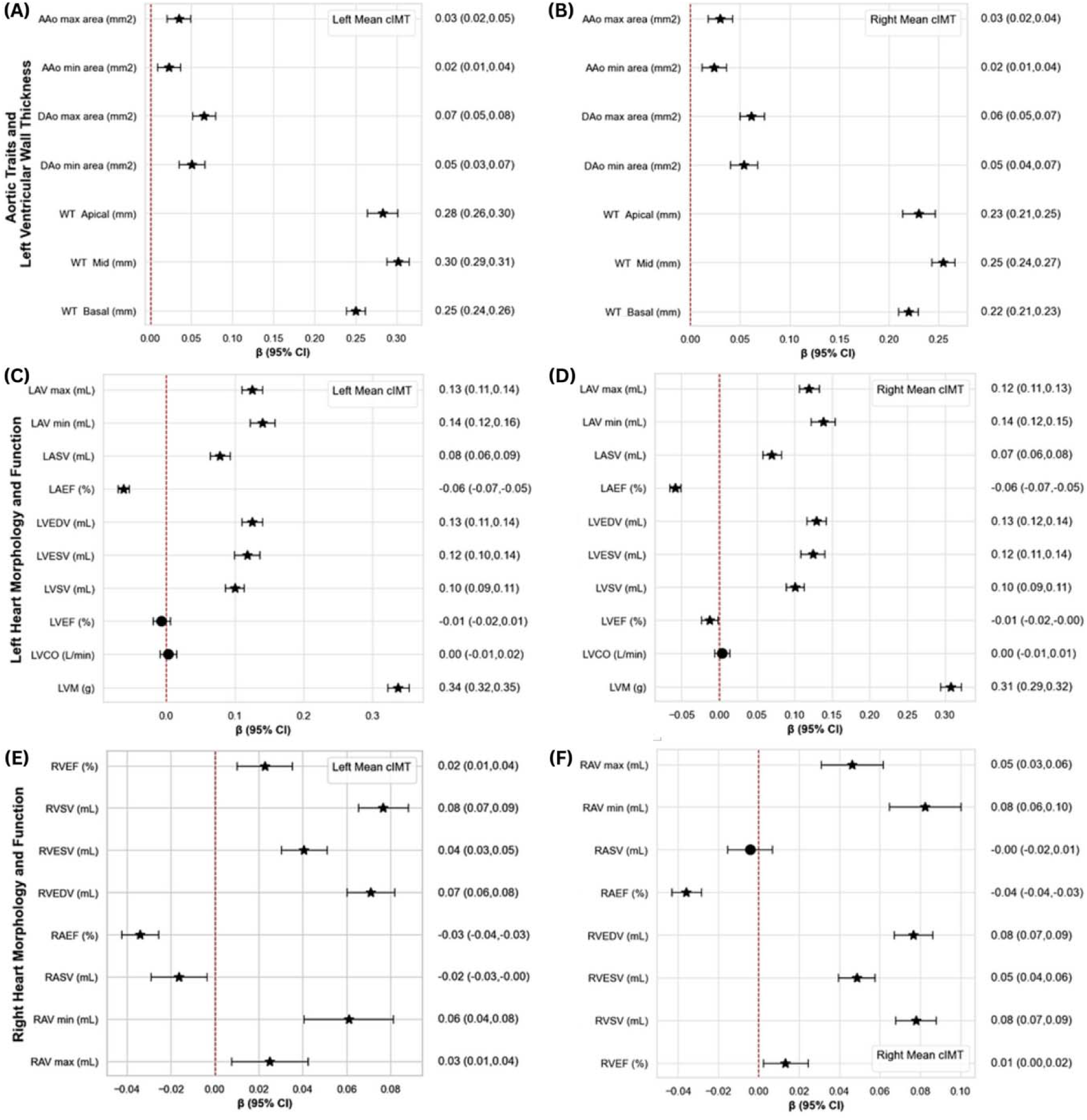
Associations between left and right mean carotid intima–media thickness (cIMT) and cardiac structure and function (N=51,818). Multivariable linear regression models examined left cIMT (A, C, E) and right cIMT (B, D, F) in relation to (A, B) aortic traits and left ventricular wall thickness, (C, D) left heart morphology and function, (E, F) right heart morphology and function. Models were adjusted for age, sex, and body mass index (BMI). Results are shown as beta coefficients (β) with corresponding 95% confidence intervals (CI). * denotes statistically significant association (p<0.05).

Higher cIMT was also associated with increased risk of AF and HF (shown in Figures 2A and 2B). In adjusted logistic regression models, elevated mean cIMT was significantly related to AF (right: P < 0.001; left: P = 0.04) and to HF for the left cIMT (P = 0.006), with a similar trend for the right side (P = 0.10) (Supplementary Table 4). No significant associations were found with cardiomyopathy, DCM, or HOCM (all P > 0.1).

**Figure 2.**
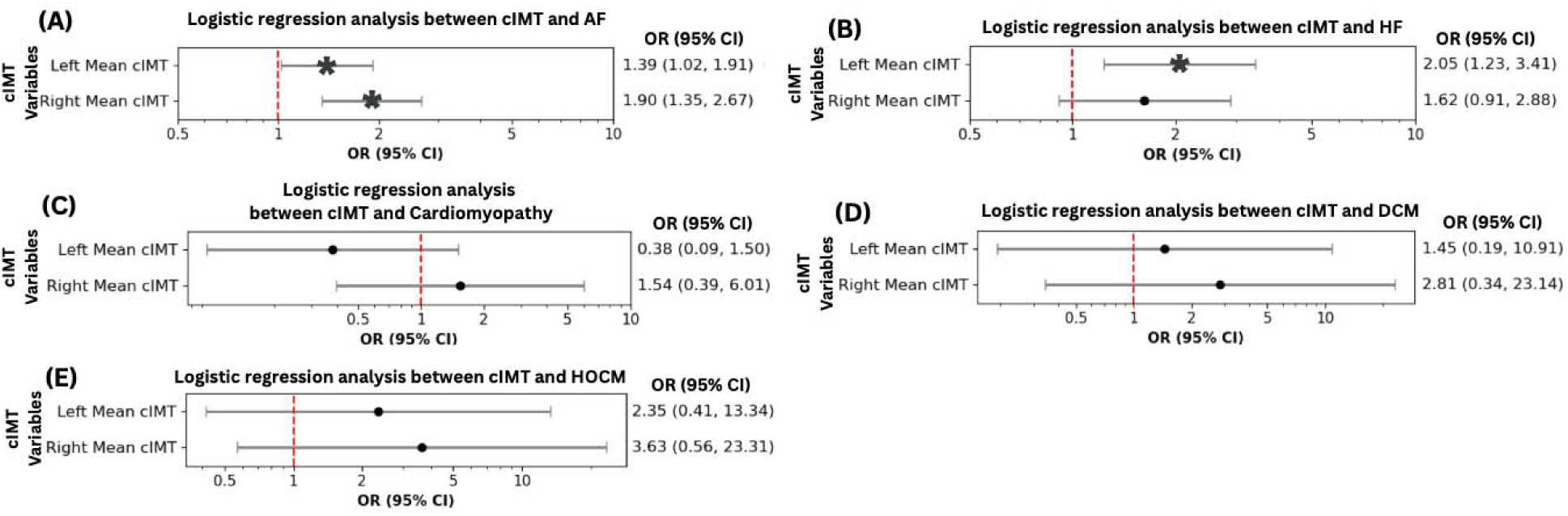
Associations between left and right mean carotid intima-media thickness (cIMT) and cardiovascular disease (CVD) (atrial fibrillation, heart failure, cardiomyopathy, dilated cardiomyopathy, hypertrophic cardiomyopathy), (N=51,818). Multivariable logistic regression models of left and right cIMT with (A) atrial fibrillation (AF), (B) heart failure (HF), (C) cardiomyopathy, (D) dilated cardiomyopathy (DCM), and (E) hypertrophic cardiomyopathy (HOCM). Models were adjusted for age, sex, and body mass index (BMI). Participants with a diagnosis of CVD prior to, or within 6 months following, the imaging visit were excluded from all analyses. Results are shown as genetic odds ratios (OR) with corresponding 95% confidence intervals (CI). * denotes statistically significant association (p<0.05).

### Genetic correlations of cIMT with aortic and cardiac phenotypes

LDSC analysis revealed significant genetic correlations between mean cIMT and measures of ventricular wall thickness, with the strongest association for LV wall thickness (WT Basal; rg = 0.50, P < 0.001), and weaker but directionally consistent correlations for WT Mid and WT Apical (Figure 3A). Positive correlations were also identified with descending aortic area (DAo maximum; rg = 0.2, P = 0.01), while no significant correlations were found with ascending aortic dimensions.

**Figure 3.**
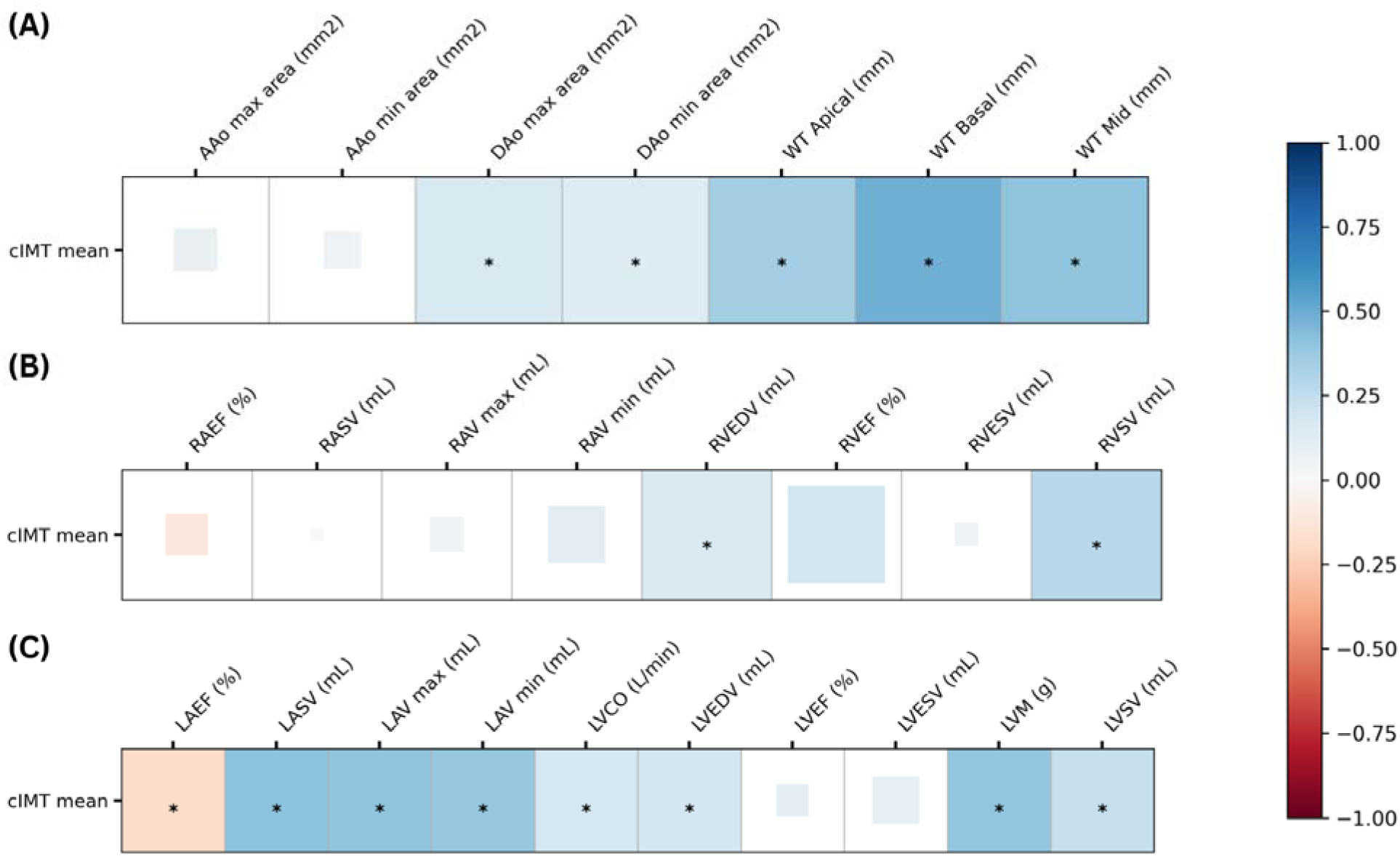
Genetic correlations between carotid intima-media thickness (cIMT) and quantitative aortic and cardiac phenotypes, (N=51,818). Linkage disequilibrium score regression (LDSC) analysis of cIMT with (A) aortic and global cardiac morphology and function, (B) right heart morphology and function, and (C) left heart morphology and function. * denotes statistically significant genetic correlations (p<0.05).

For right heart traits, cIMT showed positive genetic correlations with right ventricular end-diastolic volume (rg = 0.2, P = 0.03) and stroke volume (rg = 0.3, P < 0.001), but not with other right-sided parameters (Figure 3B). With respect to the left atrium, cIMT correlated inversely with ejection fraction (rg = –0.2, P = 0.01) and positively with atrial volume, stroke volume, and LV mass (all rg ≈ 0.4, P < 0.001) (Figure 3C); see Supplementary Table 5 for details.

### Shared genes and molecular pathways

A total of 33 independent genetic loci associated with cIMT mean, and 193 loci related to aortic and cardiac traits, as well as gene expression, were identified in the GWAS. Among the 193 prioritized genes associated with aortic and cardiac traits, seven genes, *CBFA2T3, CDH13, FBN1, FBXO32, HAND2, ITCH,* and *TBX20*, were consistently shared with cIMT mean, see supplementary Figure S2.

Pathway enrichment analysis using gProfiler highlighted biological processes central to vascular and cardiac development (Figure 4). In the Gene Ontology Molecular Function category, significant enrichment was observed for extracellular matrix (ECM) structural constituents, proteoglycan binding, and integrin interactions (adjusted *P* = 1.1×10⁻³–2.3×10⁻²). Within Biological Processes, key pathways included blood vessel and circulatory system development, vasculature morphogenesis, smooth muscle cell differentiation, and regulation of growth factor responses (*P* = 7.1×10⁻□), see supplementary Figure S3.

**Figure 4.**
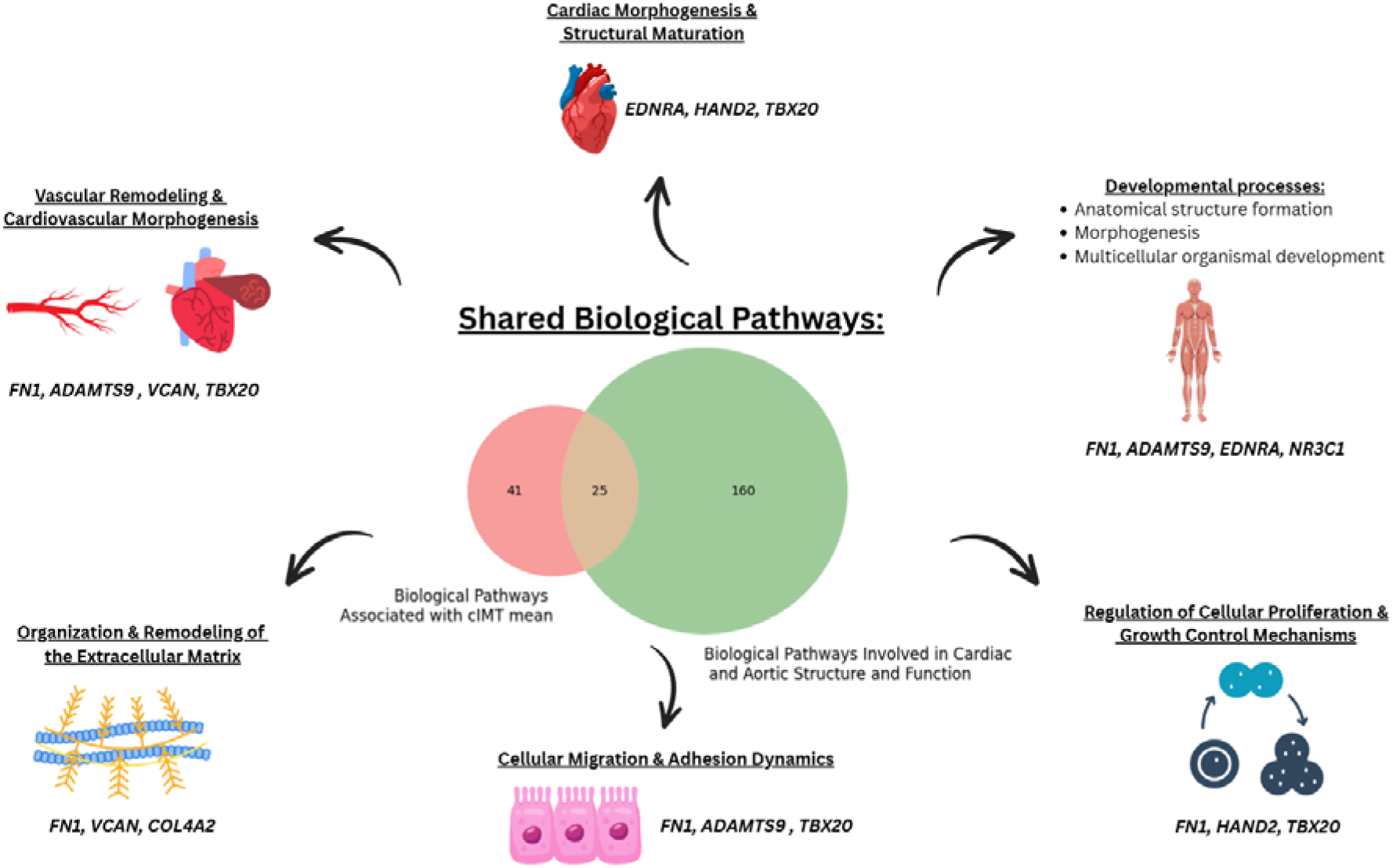
Summary of overlapping biological pathways identified for carotid intima-media thickness (cIMT), aortic, and cardiac parameters. Candidate genes associated with cIMT, aortic, and cardiac traits from the primary GWAS were analyzed for shared biological pathways and manually categorized into functional groups related to atherosclerosis and cardiac development.

Across cIMT, aortic, and cardiac traits, 25 pathways were shared (Figure 4), encompassing cardiac morphogenesis, ECM remodeling, vascular and tissue development, and cellular adhesion and migration. These convergent processes reinforce that cIMT reflects integrated vascular–cardiac remodeling mechanisms central to early CVD pathogenesis.

## DISCUSSION

To our knowledge, this is the largest population-based study to systematically investigate the relationship between cIMT, a comprehensive spectrum of aortic and cardiac phenotypes, and clinical CVD using imaging. We found that elevated cIMT was associated with subclinical structural remodeling, including increased aortic diameter, chamber enlargement, and myocardial hypertrophy, and with a higher risk of AF and HF, indicating that cIMT reflects early remodeling rather than overt systolic dysfunction. Genetic correlation analyses further revealed substantial shared heritable influences between cIMT and multiple vascular and myocardial traits, particularly descending aortic dimensions, LV wall thickness, and myocardial mass. Consistent with this, pathway analyses demonstrated convergence on biological processes involved in vascular and cardiac development, ECM organisation, and tissue remodeling, suggesting that cIMT, aortic, and cardiac traits are genetically and biologically coupled. Together, these findings position cIMT not merely as a surrogate of atherosclerosis, but as an integrated marker of systemic cardiovascular remodeling, supporting its potential utility as an early mechanistic biomarker, and a plausible future therapeutic target for interventions aimed at preventing progression to clinical CVD.

### Observational associations of cIMT, aortic and cardiac traits, and CVD

The mechanisms linking increased carotid wall thickness to downstream aortic and myocardial remodeling remain incompletely understood.^5,31–35^ Moreover, few prior studies have simultaneously examined both left and right cIMT in relation to early-stage aortic and cardiac dysfunction within large, deeply phenotyped cohorts. Consequently, incorporating data from both carotid arteries provides a more comprehensive and accurate assessment of carotid artery morphology, thereby improving the detection of subclinical vascular disease. Previous evidence suggests that cIMT reflects more than just vascular atherosclerosis. In the MESA study, cIMT independently predicted impaired myocardial strain, a marker of subclinical systolic and diastolic dysfunction, in over 500 asymptomatic individuals.^34^ Similarly, in a study of 4,301 CVD-free UK Biobank participants, higher cIMT was strongly associated with concentric remodeling of left and right ventricular left chambers, impaired atrial function, increased LV mass, and subclinical LV dysfunction.^31^

Our findings extend these observations by providing a more comprehensive characterization of atrial structural and functional changes, as well as aortic cross-sectional dimensions. We observed that higher cIMT values were associated with significantly enlarged ascending and descending aortic areas, a pattern that may reflect increased systemic vascular resistance (SVR) and altered vascular compliance. Compensatory increases in CO to maintain perfusion could impose chronic hemodynamic load on the heart, supporting the observed associations with LV wall thickening and biatrial and biventricular structural and functional alterations.^36–38^

One plausible explanation is that subclinical atherosclerosis induces myocardial ischaemia due to a supply–demand mismatch, triggering adverse remodelling, such as cardiomyocyte hypertrophy and reduced ventricular compliance.^39^ Chronic exposure to atherosclerosis-related risk factors (e.g., hypertension, dyslipidaemia, and alcohol use) may further exacerbate this process, progressively impairing myocardial structure and function.^40^ Over time, these adaptations may evolve into heart failure with preserved ejection fraction (HFpEF), or, in the presence of additional stressors such as myocardial infarction, progress to heart failure with reduced ejection fraction (HFrEF). Chronic low-grade inflammation accompanying increased carotid wall thickness may also initiate early myocardial remodeling, including hypertrophy and fibrosis, that precedes clinically overt HF.^41^

Taken together, our findings suggest that cIMT may serve not only as a marker of vascular atherosclerosis but also as an early haemodynamic biomarker of maladaptive cardiac remodeling. This dual role supports its potential utility in risk stratification and intervention before the onset of symptomatic CVD. Consistent with prior studies, such as a UK Biobank analysis of 29,292 individuals showing cIMT as an independent predictor of coronary heart disease (CHD) and MI,^9^ we also observed associations between elevated cIMT and subclinical aortic and cardiac remodeling. In contrast, we did not find significant associations with overt cardiomyopathy (dilated or hypertrophic subtypes), likely due to the predominantly healthy composition of the UK Biobank imaging cohort and limited event rates. Nonetheless, the observed associations with AF, HF, and multiple subclinical remodeling traits strongly support a continuum in which elevated cIMT precedes overt cardiovascular events such as stroke and MI. These findings reinforce the concept of cIMT as an early vascular–cardiac biomarker of systemic remodeling, rather than a late marker of atherosclerotic burden alone.

### Genetic associations of cIMT, aortic, and cardiac traits

In line with known epidemiology, the direction of genetic correlations between cIMT and traits such as ascending aortic area, right atrial chamber size and function, and LVESV was opposite to that observed in the phenotypic analysis. Nevertheless, strong positive genetic correlations were identified between cIMT and LVWT, as well as LV chamber size and volume. In contrast, LVEF showed only weak but statistically significant correlations with cIMT traits. These findings indicate that genetically mediated LV structure may play a greater role than right-sided traits in early atherosclerotic remodeling.

Previous studies have shown shared genetic correlations of cIMT with obesity and glucometabolic traits, suggesting that some variants influence vascular remodeling and metabolic regulation simultaneously.^42,43,44^ Such pathways may alter endothelial function and arterial stiffness, ultimately influencing cardiac structure. Thus, the LDSC correlations likely reflect shared heritable metabolic-vascular-cardiac mechanisms rather than secondary hemodynamic effects alone.

The divergence between genetic and observational findings likely reflects differences in study design and phenotype derivation rather than biological contradiction. Genetic approaches capture lifelong inherited influences, whereas observational analyses reflect cumulative environmental exposure, lifestyle, and comorbidity effects.^45,46,47^ Therefore, a comprehensive understanding of these relationships necessitates the integration of both inherited and modifiable risk factors. Since this is the first study to examine cIMT mean using genetic correlation analysis, independent validation is warranted.

### Commonalities and distinctions in underlying biological pathways

Only a small subset of SNPs associated with cIMT also showed associations with cardiac traits. Shared variants in genes such as *CDH13, HAND2, ITCH*, and *TBX20* align with established mechanisms in vascular remodeling, ECM dynamics, inflammation, and cardiac development, processes central to both atherosclerosis and myocardial structural changes. For example, *CDH13*, which encodes T-cadherin, is implicated in activation of the nuclear factor-κB (NF-κB) signaling pathway and smooth muscle proliferation, processes linked to carotid atherosclerosis and CVD.^48–50^ Moreover, *HAND2,* may exert protective effects by suppressing pro-inflammatory genes, thereby reducing the risk of plaque formation and cardiovascular events.^51^ *ITCH* modulates NF-κB and transforming growth factor-β (TGF-β) signaling, thereby influencing pro-atherogenic gene expression and plaque formation.^52^ In addition, *ITCH* has been implicated in cardiac hypertrophy and fibrosis through ubiquitination of key signaling molecules, contributing to both carotid atherosclerosis and pathological cardiac remodeling. Collectively, these findings suggest mechanistic links between early vascular changes and subclinical cardiac remodeling. Hence, incorporating genetic data with imaging phenotypes may enhance risk assessment and facilitate personalised preventative methods, including targeted lifestyle or pharmacological therapies to alleviate vascular and myocardial stress.

### Limitations

The study population consists predominantly of middle-aged to older individuals of White ancestry from the UK, which limits the generalizability of the findings to populations with different age distributions and ethnic backgrounds. Moreover, the cross-sectional study design precludes the establishment of causal inferences regarding the observed relationships between carotid atherosclerosis, aortic, and cardia traits. Additionally, although multivariable analyses were adjusted for key potential confounders, the influence of residual or unmeasured confounding factors cannot be entirely ruled out and may contribute to the observed associations. In the genetic analysis, although shared genes were identified, limitations include polygenicity, measurement variability, and heterogeneity across phenotypes, which may have reduced detectable associations. Future studies should incorporate longitudinal imaging and multi-omics to elucidate how variants contribute to cardiovascular remodeling over time.

## CONCLUSION

We demonstrated that elevated cIMT was associated with alterations in cardiac structure and function and with a higher prevalence of CVD, particularly AF and HF. Genetic correlation analyses further revealed that cIMT, aortic dimensions, and cardiac traits share common genetic architecture and biological pathways, including those involved in aortic and cardiac chamber development, extracellular matrix organisation, and cellular proliferation. These findings suggest that routine cIMT assessment in high-risk individuals may enable earlier identification of subclinical cardiac remodelling before overt CVD becomes clinically manifest, supporting its potential utility as an early, non-invasive biomarker for risk stratification, surveillance, and preventive intervention. The results reinforce the need for comprehensive management of cardiovascular risk factors and motivate future investigation into the transcriptomic and epigenetic mechanisms driving these associations.

## DATA AVAILABILITY STATEMENT

This research was conducted using data from the UK Biobank (Application Number 12113). The data used in this study are available to bona fide researchers upon application. Derived data generated in this study will be returned to the UK Biobank in accordance with their data sharing policy and will be made available to future approved researchers through the UK Biobank resource.

## AUTHOR CONTRIBUTIONS

SHC was responsible for data curation, formal analysis, investigation, and drafting of the manuscript. AS, MV were responsible for data acquisition and contributed to data sharing. MV, AS, and AFS contributed to reviewing and editing the manuscript and validation of the analyses. NP led the conceptualization and methodology, supervised the project, and contributed to reviewing and editing the manuscript. All authors have read and approved the final version of the manuscript.

## REFERENCES

1. Gisterå A, Hansson GK. The immunology of atherosclerosis. Nature reviews nephrology. 2017;13(6):368–380.

2. Zhu Y, Xian X, Wang Z, et al. Research Progress on the Relationship between Atherosclerosis and Inflammation. Biomolecules. 2018;8(3).

3. Strawbridge RJ, Ward J, Bailey MES, et al. Carotid Intima-Media Thickness: Novel Loci, Sex-Specific Effects, and Genetic Correlations With Obesity and Glucometabolic Traits in UK Biobank. Arterioscler Thromb Vasc Biol. 2020;40(2):446–461.

4. O’Leary DH, Polak JF, Kronmal RA, Manolio TA, Burke GL, Wolfson SK, Jr. Carotid-artery intima and media thickness as a risk factor for myocardial infarction and stroke in older adults. Cardiovascular Health Study Collaborative Research Group. N Engl J Med. 1999;340(1):14–22.

5. Mitra S, Biswas RK, Hooijenga P, et al. Carotid intima-media thickness, cardiovascular disease, and risk factors in 29,000 UK Biobank adults. American Journal of Preventive Cardiology. 2025;22:101011.

6. Yeboah J, McClelland RL, Polonsky TS, et al. Comparison of novel risk markers for improvement in cardiovascular risk assessment in intermediate-risk individuals. Jama. 2012;308(8):788–795.

7. Nambi V, Chambless L, Folsom AR, et al. Carotid intima-media thickness and presence or absence of plaque improves prediction of coronary heart disease risk: the ARIC (Atherosclerosis Risk In Communities) study. J Am Coll Cardiol. 2010;55(15):1600–1607.

8. Bulik-Sullivan B, Finucane HK, Anttila V, et al. An atlas of genetic correlations across human diseases and traits. Nat Genet. 2015;47(11):1236–1241.

9. Mitra S, Biswas RK, Hooijenga P, et al. Carotid intima-media thickness, cardiovascular disease, and risk factors in 29,000 UK Biobank adults. Am J Prev Cardiol. 2025;22:101011.

10. McGill Jr HC, McMahan CA, Herderick EE, et al. Origin of atherosclerosis in childhood and adolescence. The American journal of clinical nutrition. 2000;72(5):1307s–1315s.

11. Bis JC, Kavousi M, Franceschini N, et al. Meta-analysis of genome-wide association studies from the CHARGE consortium identifies common variants associated with carotid intima media thickness and plaque. Nature genetics. 2011;43(10):940–947.

12. Natarajan P, Bis JC, Bielak LF, et al. Multiethnic exome-wide association study of subclinical atherosclerosis. Circulation: Cardiovascular Genetics. 2016;9(6):511–520.

13. Franceschini N, Giambartolomei C, De Vries PS, et al. GWAS and colocalization analyses implicate carotid intima-media thickness and carotid plaque loci in cardiovascular outcomes. Nature communications. 2018;9(1):5141.

14. Lakda S, Davies RH, Hingorani AD, Paliwal N. A systematic review of GWAS on CMR imaging traits: genetic insights into cardiovascular structure, function, and diseases. EBioMedicine. 2025;121:105992.

15. Psaty BM, O’Donnell CJ, Gudnason V, et al. Cohorts for Heart and Aging Research in Genomic Epidemiology (CHARGE) Consortium. Circulation: Cardiovascular Genetics. 2009;2(1):73–80.

16. Sudlow C, Gallacher J, Allen N, et al. UK biobank: an open access resource for identifying the causes of a wide range of complex diseases of middle and old age. PLoS medicine. 2015;12(3):e1001779.

17. Littlejohns TJ, Holliday J, Gibson LM, et al. The UK Biobank imaging enhancement of 100,000 participants: rationale, data collection, management and future directions. Nature Communications. 2020;11(1):2624.

18. Petersen SE, Matthews PM, Bamberg F, et al. Imaging in population science: cardiovascular magnetic resonance in 100,000 participants of UK Biobank-rationale, challenges and approaches. Journal of Cardiovascular Magnetic Resonance. 2013;15(1):46.

19. Coffey S, Lewandowski AJ, Garratt S, et al. Protocol and quality assurance for carotid imaging in 100,000 participants of UK Biobank: development and assessment. European journal of preventive cardiology. 2017;24(17):1799–1806.

20. Peters S, Lind L, Palmer M, et al. Increased age, high body mass index and low HDL-C levels are related to an echolucent carotid intima–media: the METEOR study. Journal of internal medicine. 2012;272(3):257–266.

21. Breuel TM. High Performance Text Recognition Using a Hybrid Convolutional-LSTM Implementation. Paper presented at: 2017 14th IAPR International Conference on Document Analysis and Recognition (ICDAR); 9-15 Nov. 2017, 2017.

22. Petersen SE, Matthews PM, Francis JM, et al. UK Biobank’s cardiovascular magnetic resonance protocol. Journal of cardiovascular magnetic resonance. 2016;18(1):8.

23. Petersen SE, Aung N, Sanghvi MM, et al. Reference ranges for cardiac structure and function using cardiovascular magnetic resonance (CMR) in Caucasians from the UK Biobank population cohort. Journal of cardiovascular magnetic resonance. 2016;19(1):18.

24. Petersen SE, Matthews PM, Francis JM, et al. UK Biobank’s cardiovascular magnetic resonance protocol. J Cardiovasc Magn Reson. 2016;18:8.

25. Bai W, Sinclair M, Tarroni G, et al. Automated cardiovascular magnetic resonance image analysis with fully convolutional networks. Journal of Cardiovascular Magnetic Resonance. 2018;20(1):65.

26. Paliwal N, Henry A, Finan C, et al. Abstract 4141107: Genome-wide association study of aortic stiffness derived from deep learning in CMR images of 45,789 individuals identifies loci linked with cell-matrix structure. Circulation. 2024;150(Suppl_1):A4141107–A4141107.

27. Bulik-Sullivan B, Finucane HK, Anttila V, et al. An atlas of genetic correlations across human diseases and traits. Nature genetics. 2015;47(11):1236–1241.

28. Wang M, Wang L, Ghazal R, et al. Abstract 4141605: Genetic Determinant for Prognosis of Dilated Cardiomyopathy: The role of NAV3 in Cardiac Fibrosis. Circulation. 2024;150(Suppl_1):A4141605–A4141605.

29. He Y, Masaru Koido, Yuka Shimmori, and Yoichiro Kamatani. GWASLab: A Python Package for Processing and Visualizing GWAS Summary Statistics. 2015.

30. Aung N, Vargas JD, Yang C, et al. Genome-wide association analysis reveals insights into the genetic architecture of right ventricular structure and function. Nat Genet. 2022;54(6):783–791.

31. Simon J, Fung K, Raisi-Estabragh Z, et al. Association between subclinical atherosclerosis and cardiac structure and function—results from the UK Biobank Study. European Heart Journal - Imaging Methods and Practice. 2023;1(2).

32. Yeung MW, Wang S, van de Vegte YJ, et al. Twenty-Five Novel Loci for Carotid Intima-Media Thickness: A Genome-Wide Association Study in >45 000 Individuals and Meta-Analysis of >100 000 Individuals. Arterioscler Thromb Vasc Biol. 2022;42(4):484–501.

33. Robert C, Tan WY, Ling L-H, Hilal S. Association of arterial structure and function with incident cardiovascular diseases and cognitive decline. *Alzheimer’s & Dementia: Diagnosis*, Assessment & Disease Monitoring. 2025;17(1):e70069.

34. Fernandes VRS, Polak JF, Edvardsen T, et al. Subclinical Atherosclerosis and Incipient Regional Myocardial Dysfunction in Asymptomatic Individuals: The Multi-Ethnic Study of Atherosclerosis (MESA). Journal of the American College of Cardiology. 2006;47(12):2420–2428.

35. Deak JD. Using Local and Global Genetic Correlation Approaches to Help Elucidate the Shared Genetic Etiology of Psychiatric and Substance Use Traits. Biol Psychiatry. 2022;92(7):e31–e33.

36. Pirruccello JP, Chaffin MD, Chou EL, et al. Deep learning enables genetic analysis of the human thoracic aorta. Nat Genet. 2022;54(1):40–51.

37. Rizzoni D, Muiesan ML, Porteri E, et al. Relations between cardiac and vascular structure in patients with primary and secondary hypertension. Journal of the American College of Cardiology. 1998;32(4):985–992.

38. Sorof JM, Alexandrov AV, Cardwell G, Portman RJ. Carotid artery intimal-medial thickness and left ventricular hypertrophy in children with elevated blood pressure. Pediatrics. 2003;111(1):61–66.

39. Kallikazaros I, Tsioufis C, Sideris S, Stefanadis C, Toutouzas P. Carotid artery disease as a marker for the presence of severe coronary artery disease in patients evaluated for chest pain. Stroke. 1999;30(5):1002–1007.

40. Drazner MH. The progression of hypertensive heart disease. Circulation. 2011;123(3):327–334.

41. Torre-Amione G, Kapadia S, Benedict C, Oral H, Young JB, Mann DL. Proinflammatory cytokine levels in patients with depressed left ventricular ejection fraction: a report from the Studies of Left Ventricular Dysfunction (SOLVD). J Am Coll Cardiol. 1996;27(5):1201–1206.

42. Franceschini N, Giambartolomei C, de Vries PS, et al. GWAS and colocalization analyses implicate carotid intima-media thickness and carotid plaque loci in cardiovascular outcomes. Nature Communications. 2018;9(1):5141.

43. Strawbridge RJ, Ward J, Bailey MES, et al. Carotid Intima-Media Thickness. Arteriosclerosis, Thrombosis, and Vascular Biology. 2020;40(2):446–461.

44. Doliner B, Dong C, Blanton SH, et al. Apolipoprotein E Gene Polymorphism and Subclinical Carotid Atherosclerosis: The Northern Manhattan Study. J Stroke Cerebrovasc Dis. 2018;27(3):645–652.

45. Uffelmann E, Huang QQ, Munung NS, et al. Genome-wide association studies. Nature Reviews Methods Primers. 2021;1(1):59.

46. Schoeler T, Speed D, Porcu E, Pirastu N, Pingault J-B, Kutalik Z. Participation bias in the UK Biobank distorts genetic associations and downstream analyses. Nature Human Behaviour. 2023;7(7):1216–1227.

47. Zhang Y, Cheng Y, Jiang W, Ye Y, Lu Q, Zhao H. Comparison of methods for estimating genetic correlation between complex traits using GWAS summary statistics. Brief Bioinform. 2021;22(5).

48. Lee JH, Shin DJ, Park S, Kang SM, Jang Y, Lee SH. Association between CDH13 variants and cardiometabolic and vascular phenotypes in a Korean population. Yonsei Med J. 2013;54(6):1305–1312.

49. Wen Y, Ma L, Liu Y, Xiong H, Shi D. Decoding the enigmatic role of T-cadherin in tumor angiogenesis. Front Immunol. 2025;16:1564130.

50. Rubina KA, Semina EV, Kalinina NI, Sysoeva VY, Balatskiy AV, Tkachuk VA. Revisiting the multiple roles of T-cadherin in health and disease. Eur J Cell Biol. 2021;100(7-8):151183.

51. Tsuchihashi T, Maeda J, Shin CH, et al. Hand2 function in second heart field progenitors is essential for cardiogenesis. Dev Biol. 2011;351(1):62–69.

52. Deng Z, Fan T, Xiao C, et al. TGF-β signaling in health, disease, and therapeutics. Signal Transduct Target Ther. 2024;9(1):61.

