## Supplementary Tables and Figures for "Integrative observational and genetic evidence from the UK Biobank supports carotid intima-media thickness as a window into cardiac remodeling"

### **SUPPLEMENTARY MATERIAL**

Supplementary Table 1: Aortic and cardiac traits included in the study. Wall thickness measures include apical, mid, and basal segments. Right heart traits comprise volumes and ejection fraction of the right atrium and ventricle, including maximal and minimal volumes and stroke volumes. Left heart traits include analogous measures of the left atrium and ventricle, along with left ventricular cardiac output and mass. The main text provides a summary of the number of participants for whom data is available for each characteristic. AAo: Ascending aorta; DAo: Descending aorta; WT apical: wall thickness at apical of the heart; WT Mid: wall thickness at mid-ventricular of the heart; WT basal: wall thickness at the base of the heart; RAV: Right Atrial Volume; RASV: Right Atrial Stroke Volume; RAEF: Right Atrial Ejection Fraction; RVEDV: Right Ventricular End-Diastolic Volume; RVESV: Right Ventricular End Systolic Volume; RVSV: Right Ventricular Stroke Volume; RVEF: Right Ventricular Ejection Fraction; LAV: Left Atrial Volume; LASV: Left Atrial Stroke Volume; LAEF: Left Atrial Ejection Fraction; LVEDV: Left Ventricular End-Diastolic Volume; LVESV: Left Ventricular End Systolic Volume; LVSV: Left Ventricular Stroke Volume; LVEF: Left Ventricular Ejection Fraction; LVCO: Left Ventricular Cardiac Output; LVM: Left Ventricular Mass

| **Aortic** | **Left Heart** | **Right Heart** | **Left Ventricular Structure** |
| --- | --- | --- | --- |
| AAo max area (mm2) | LAV max (mL) | RAV max (mL) | WT Apical (mm) |
| AAo min area (mm2) | LAV min (mL) | RAV min (mL) | WT Mid (mm) |
| DAo max area (mm2) | LASV (mL) | RASV (mL) | WT Basal (mm) |
| DAo min area (mm2) | LAEF (%) | RAEF (%) | LVCO (L/min) |
|  | LVEDV (mL) | RVEDV (mL) | LVM (g) |
|  | LVESV (mL) | RVESV (mL) |  |
|  | LVSV (mL) | RVSV (mL) |  |
|  | LVEF (%) | RVEF (%) |  |

Supplementary Table 2: carotid intima-media thickness (cIMT), aortic and cardiac shape and function of the UK Biobank participants stratified by sex. Data are reported as mean ± SD for normal distribution, non-normal distribution data are expressed as number, median (Q1, Q3), Categorial variables are summarised as %. cIMT: Carotid intima-media thickness; AAo: Ascending aorta; DAo: Decending aorta; WT apical: wall thickness at apical of the heart; WT Mid: wall thickness at mid-ventricular of the heart; WT basal: wall thickness at the base of the heart; RAV: Right Atrial Volume; RASV: Right Atrial Stroke Volume; RAEF: Right Atrial Ejection Fraction; RVEDV: Right Ventricular End-Diastolic Volume; RVESV: Right Ventricular End Systolic Volume; RVSV: Right Ventricular Stroke Volume; RVEF: Right Ventricular Ejection Fraction; LAV: Left Atrial Volume; LASV: Left Atrial Stroke Volume; LAEF: Left Atrial Ejection Fraction; LVEDV: Left Ventricular End-Diastolic Volume; LVESV: Left Ventricular End Systolic Volume; LVSV: Left Ventricular Stroke Volume; LVEF: Left Ventricular Ejection Fraction; LVCO: Left Ventricular Cardiac Output; LVM: Left Ventricular Mass.

| Cardiac traits | Overall (N=51,818) | Female  (N=26,533) | Male  (N=25,285) |
| --- | --- | --- | --- |
| Aorta |  |  |  |
| AAo max area (mm2) | 839.83 (727.01, 972.69) | 769.63 (676.88, 879.94) | 920.05 (804.73, 1047.90) |
| AAo min area (mm2) | 764.62 (654.31, 894.98) | 701.94 (606.68, 809.74) | 839.83 (727.01, 965.17) |
| DAo max area (mm2) | 468.80 (408.63, 538.99) | 418.66 (376.04, 468.80) | 526.46 (471.31, 589.13) |
| DAo min area (mm2) | 413.65 (355.99, 481.33) | 368.52 (325.90, 416.15) | 468.80 (416.15, 528.97) |
| Left Ventricular Wall Thickness (WT) |  |  |  |
| WT Apical (mm) | 18.52 (16.57, 20.64) | 16.88 (15.49, 18.32) | 20.44 (18.84, 22.07) |
| WT Mid (mm) | 35.26 (31.76, 39.07) | 32.09 (29.91, 34.52) | 38.74 (36.14, 41.68) |
| WT Basal (mm) | 35.28 (31.94, 39.07) | 32.68 (30.22, 35.40) | 38.38 (35.31, 41.59) |
| Right Heart Structure and Function |  |  |  |
| RAV max (mL) | 82.14 (67.31, 101.27) | 73.66 (61.94, 87.17) | 94.35 (76.65, 114.43) |
| RAV min (mL) | 42.90 (33.26, 55.53) | 36.97 (29.71, 45.27) | 51.69 (40.45, 65.10) |
| RASV (mL) | 38.42 (30.88, 47.55) | 36.11 (29.38, 43.85) | 41.45 (33.01, 51.33) |
| RAEF | 46.89 (41.12, 52.89) | 49.29 (43.78, 54.93) | 44.33 (38.87, 50.13) |
| RVEDV (mL) | 151.56 (128.37, 179.79) | 131.63 (116.59, 147.79) | 177.84 (156.77, 200.76) |
| RVESV (mL) | 64.35 (51.57, 80.39) | 53.28 (45.19, 62.48) | 79.14 (67.15, 92.76) |
| RVSV (mL) | 86.71 (74.33, 101.40) | 78.10 (68.66, 88.01) | 98.43 (85.73, 111.78) |
| RVEF | 57.54 (53.51, 61.34) | 59.45 (55.80, 62.97) | 55.47 (51.61, 59.11) |
| Left Heart Structure and Function |  |  |  |
| LAV max (mL) | 69.66 (56.11, 85.51) | 65.44 (53.34, 78.34) | 75.52 (60.08, 92.94) |
| LAV min (mL) | 27.03 (19.57, 35.91) | 24.98 (18.49, 32.48) | 29.69 (21.01, 39.72) |
| LASV (mL) | 42.01 (34.77, 50.22) | 39.89 (33.42, 46.79) | 44.93 (36.64, 53.68) |
| LAEF | 60.97 (55.80, 66.39) | 61.47 (56.58, 66.68) | 60.45 (54.98, 66.07) |
| LVEDV (mL) | 142.78 (122.34, 167.53) | 126.20 (112.21, 141.68) | 164.63 (145.25, 185.57) |
| LVESV (mL) | 56.91 (46.26, 70.52) | 48.30 (41.19, 56.57) | 68.59 (58.01, 81.20) |
| LVSV (mL) | 84.91 (73.26, 98.56) | 77.13 (68.02, 87.28) | 94.82 (82.87, 107.94) |
| LVEF | 59.83 (55.86, 63.77) | 61.43 (57.76, 65.10) | 58.02 (54.08, 61.98) |
| LVCO (L/min) | 5.26 (4.53, 6.12) | 4.83 (4.22, 5.57) | 5.73 (5.02, 6.57) |
| LVM (g) | 82.63 (68.41, 100.37) | 69.10 (61.93, 77.27) | 100.09 (89.26, 112.35) |

Supplementary Table 3: Multivariable linear regression analysis examining the association between carotid intima-media thickness (cIMT) and aortic, as well as cardiac traits among UK Biobank participants, adjusted for age, sex, and BMI*.* B: Beta Coefficient; CI: Confidence Interval; AAo: Ascending aorta; DAo: Descending aorta; WT apical: wall thickness at apical of the heart; WT Mid: wall thickness at mid-ventricular of the heart; WT basal: wall thickness at the base of the heart; RAV: Right Atrial Volume; RASV: Right Atrial Stroke Volume; RAEF: Right Atrial Ejection Fraction; RVEDV: Right Ventricular End-Diastolic Volume; RVESV: Right Ventricular End Systolic Volume; RVSV: Right Ventricular Stroke Volume; RVEF: Right Ventricular Ejection Fraction; LAV: Left Atrial Volume; LASV: Left Atrial Stroke Volume; LAEF: Left Atrial Ejection Fraction; LVEDV: Left Ventricular End-Diastolic Volume; LVESV: Left Ventricular End Systolic Volume; LVSV: Left Ventricular Stroke Volume; LVEF: Left Ventricular Ejection Fraction; LVCO: Left Ventricular Cardiac Output; LVM: Left Ventricular Mass

| **Continuous Variables** | **UK Biobank Participants**  **N=51,818** | | | |
| --- | --- | --- | --- | --- |
|  | **x= Right Mean, y= Cardiac Traits** | | **x= Left Mean, y= Cardiac Traits** | |
|  | **B (95% CI)** | **P-value** | **B (95% CI)** | **P-value** |
| **Aortic Traits** |  |  |  |  |
| AAo max area(mm2) | 0.03 (0.018, 0.043) | <0.001 | 0.035 (0.021, 0.049) | <0.001 |
| AAo min area (mm2) | 0.024 (0.012, 0.036) | <0.001 | 0.023 (0.009, 0.037) | <0.001 |
| DAo max area (mm2) | 0.062 (0.050, 0.074) | <0.001 | 0.065 (0.051, 0.079) | <0.001 |
| DAo min area (mm2) | 0.054 (0.040, 0.067) | <0.001 | 0.050 (0.035, 0.066) | <0.001 |
| **Left Ventricular Wall Thickness (WT)** |  |  |  |  |
| WT Apical (mm) | 0.230 (0.214, 0.246) | <0.001 | 0.283 (0.264, 0.30) | <0.001 |
| WT Mid (mm) | 0.254 (0.243, 0.266) | <0.001 | 0.302 (0.288, 0.315) | <0.001 |
| WT Basal (mm) | 0.219 (0.209, 0.229) | <0.001 | 0.250 (0.238, 0.261) | <0.001 |
| **Right Heart Structure and Function** |  |  |  |  |
| RAV max (mL) | 0.046 (0.031, 0.061) | <0.001 | 0.025 (0.008, 0.043) | 0.057 |
| RAV min (mL) | 0.082 (0.065, 0.100) | <0.001 | 0.061 (0.041, 0.081) | <0.001 |
| RASV (mL) | -0.004 (-0.016, 0.007) | 0.447 | -0.016 (-0.029, -0.004) | 0.007 |
| RAEF (%) | -0.036 (-0.043, -0.029) | <0.001 | -0.034 (-0.043, -0.026) | <0.001 |
| RVEDV (mL) | 0.077 (0.067, 0.086) | <0.001 | 0.071 (0.060, 0.082) | <0.001 |
| RVESV (mL) | 0.049 (0.040, 0.057) | <0.001 | 0.041 (0.030, 0.051) | <0.001 |
| RVSV (mL) | 0.079 (0.068, 0.088) | <0.001 | 0.077 (0.065, 0.088) | <0.001 |
| RVEF (%) | 0.013 (0.002, 0.024) | 0.018 | 0.023 (0.010, 0.035) | <0.001 |
| **Left Heart Structure and Function** |  |  |  |  |
| LAV max (mL) | 0.119 (0.106, 0.133) | <0.001 | 0.125 (0.110, 0.141) | <0.001 |
| LAV min (mL) | 0.138 (0.122, 0.154) | <0.001 | 0.140 (0.122, 0.158) | <0.001 |
| LASV (mL) | 0.07 (0.058, 0.082) | <0.001 | 0.079 (0.064, 0.093) | <0.001 |
| LAEF (%) | -0.059 (-0.066, -0.052) | <0.001 | -0.062 (-0.070, -0.054) | <0.001 |
| LVEDV (mL) | 0.129 (0.116, 0.142) | <0.001 | 0.125 (0.111, 0.140) | <0.001 |
| LVESV (mL) | 0.124 (0.108, 0.140) | <0.001 | 0.118 (0.099, 0.136) | <0.001 |
| LVSV (mL) | 0.100 (0.089, 0.112) | <0.001 | 0.099 (0.087, 0.11) | <0.001 |
| LVEF (%) | -0.013 (-0.023, -0.002) | 0.065 | -0.006 (-0.019, 0.006) | 0.364 |
| LVCO (L/min) | 0.0039 (-0.0064, 0.0142) | 0.4567 | 0.003 (-0.008, 0.015) | 0.69 |
| LVM (g) | 0.3079 (0.2945, 0.3214) | <0.001 | 0.337 (0.322, 0.353) | <0.001 |

Supplementary Table 4: Multivariable logistic regression analysis examining the association between carotid intima-media thickness (cIMT) and cardiovascular disease (CVD) among UK Biobank participants, adjusted for age, sex, and body mass index (BMI)*.* OR: Odd Ratio; CI: Confidence Interval

| **Binary Variables** | **UK Biobank Participants**  **N=51,818** | | | |
| --- | --- | --- | --- | --- |
|  | **x= Right Mean, y= CVD** | | **x= Left Mean, y= CVD** | |
|  | **OR (95% CI)** | **P-value** | **OR (95% CI)** | **P-value** |
| Atrial Fibrillation | 1.899 (1.349, 2.673) | <0.001 | 1.392 (1.016, 1.907) | 0.040 |
| Heart Failure | 1.619 (0.909, 2.883) | 0.102 | 2.052 (1.235, 3.411) | 0.006 |
| Cardiomyopathy | 1.540 (0.394, 6.015) | 0.534 | 0.377 (0.094, 1.505) | 0.167 |
| Dilated Cardiomyopathy | 2.81 (0.341, 23.139) | 0.337 | 1.448 (0.192, 10.91) | 0.720 |
| Hypertrophic Cardiomyopathy | 3.629 (0.565, 23.310) | 0.174 | 2.348 (0.413, 13.343) | 0.336 |

Supplementary Table 5: Genetic correlation between carotid intima-media thickness (cIMT), aortic, and cardiac traits using linkage disequilibrium (LD) score. Rg: Genetic correlation; CI: Confidence Interval; AAo: ascending aorta; DAo: descending aorta; WT apical: wall thickness at apical of the heart; WT Mid: wall thickness at mid-ventricular of the heart; WT basal: wall thickness at the base of the heart; RAV: Right Atrial Volume; RASV: Right Atrial Stroke Volume; RAEF: Right Atrial Ejection Fraction; RVEDV: Right Ventricular End-Diastolic Volume; RVESV: Right Ventricular End Systolic Volume; RVSV: Right Ventricular Stroke Volume; RVEF: Right Ventricular Ejection Fraction; LAV: Left Atrial Volume; LASV: Left Atrial Stroke Volume; LAEF: Left Atrial Ejection Fraction; LVEDV: Left Ventricular End-Diastolic Volume; LVESV: Left Ventricular End Systolic Volume; LVSV: Left Ventricular Stroke Volume; LVEF: Left Ventricular Ejection Fraction; LVCO: Left Ventricular Cardiac Output; LVM: Left Ventricular Mass

| **Continuous Variables** | **cIMT Mean** | |
| --- | --- | --- |
|  | **Rg (95% CI)** | **P-value** |
| **Aortic Traits** |  |  |
| AAo max area(mm2) | 0.08 (-0.11, 0.26) | 0.41 |
| AAo min area (mm2) | 0.06 (-0.11, 0.23) | 0.47 |
| DAo max area (mm2) | 0.17 (0.04, 0.29) | 0.01 |
| DAo min area (mm2) | 0.14 (0.01, 0.27) | 0.04 |
| **Left Ventricular Wall Thickness (WT)** |  |  |
| WT Apical (mm) | 0.35 (0.24, 0.45) | <0.001 |
| WT Mid (mm) | 0.40 (0.29, 0.51) | <0.001 |
| WT Basal (mm) | 0.49 (0.39, 0.58) | <0.001 |
| **Right Heart Structure and Function** |  |  |
| RAV max (mL) | 0.04 (-0.07, 0.16) | 0.45 |
| RAV min (mL) | 0.10 (-0.08, 0.28) | 0.27 |
| RASV (mL) | 0.01 (-0.12, 0.13) | 0.91 |
| RAEF (%) | -0.11 (-0.35, 0.13) | 0.38 |
| RVEDV (mL) | 0.16 (0.04, 0.28) | 0.01 |
| RVESV (mL) | 0.05 (-0.13, 0.22) | 0.58 |
| RVSV (mL) | 0.28 (0.16, 0.40) | <0.001 |
| RVEF (%) | 0.19 (-0.04, 0.42) | 0.11 |
| **Left Heart Structure and Function** |  |  |
| LAV max (mL) | 0.40 (0.25, 0.55) | <0.001 |
| LAV min (mL) | 0.38 (0.26, 0.51) | <0.001 |
| LASV (mL) | 0.41 (0.23, 0.59) | <0.001 |
| LAEF (%) | -0.19 (-0.34, -0.04) | 0.01 |
| LVEDV (mL) | 0.18 (0.08, 0.29) | <0.001 |
| LVESV (mL) | 0.09 (-0.07, 0.25) | 0.25 |
| LVSV (mL) | 0.24 (0.13, 0.35) | <0.001 |
| LVEF (%) | 0.10 (-0.12, 0.31) | 0.39 |
| LVCO (L/min) | 0.17 (0.03, 0.32) | 0.02 |
| LVM (g) | 0.39 (0.31, 0.47) | <0.001 |

Supplementary Figure S1: Representative carotid ultrasound report showing B-mode images of the left and right common carotid arteries with corresponding cIMT measurements.

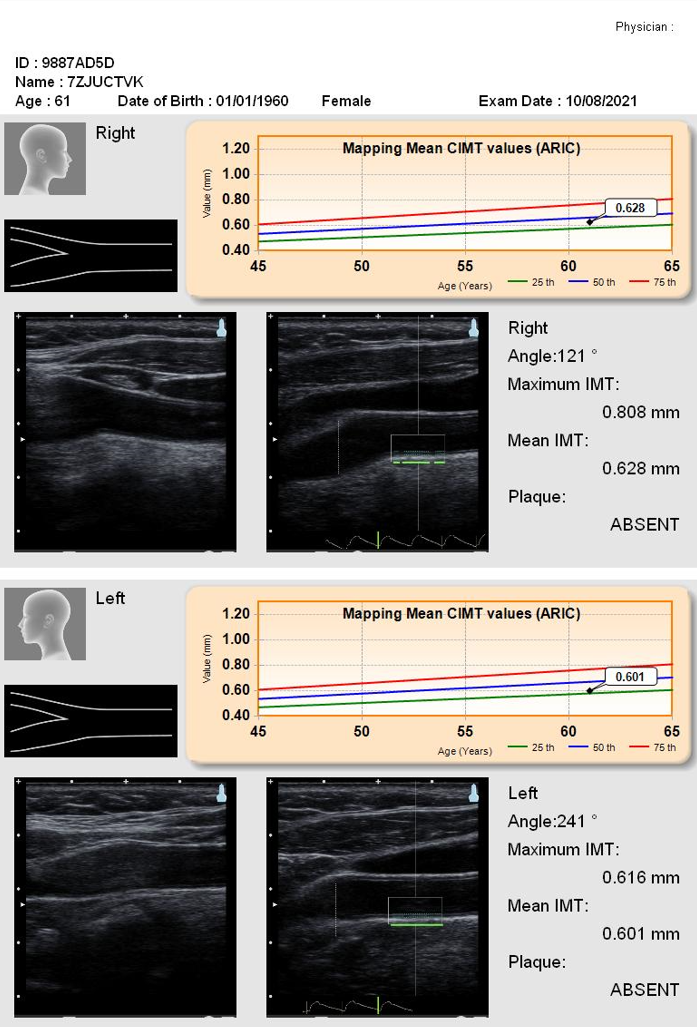

Supplementary Figure S2: Summary of independent and overlapping genetic loci identified for carotid intima-media thickness (cIMT), aortic, and cardiac parameters Venn diagram illustrates the overlap of independent loci identified through GWAS of cIMT phenotype and a comprehensive set of aortic and cardiac traits. Aortic and cardiac loci include both structural and functional parameters of the heart and great vessels. Shared and trait-specific loci are annotated with the nearest protein-coding gene to the lead variant.

.

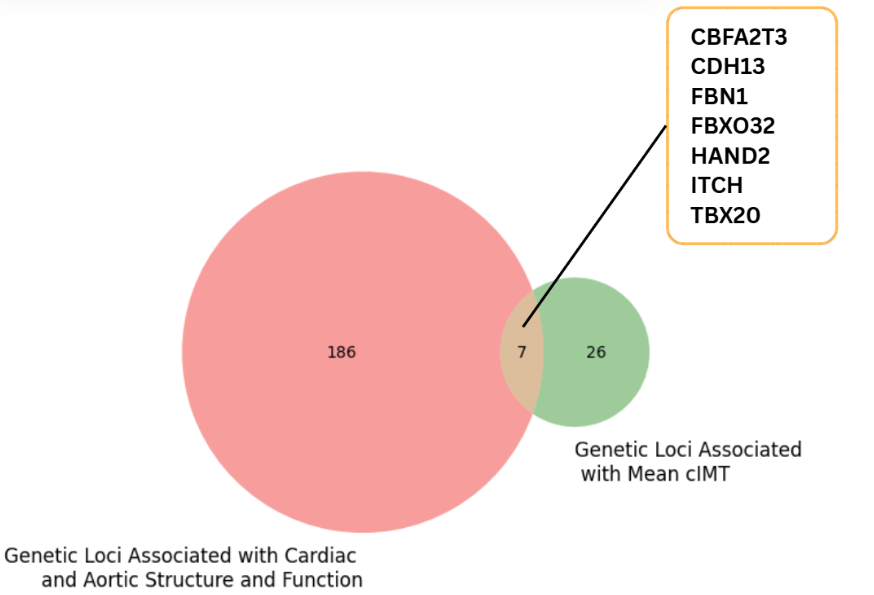

Supplementary Figure S3: Biological pathway analysis of mean carotid intima-media thickness (cIMT) using gProfiler. Significantly enriched biological pathways identified for genetic loci associated with cIMT mean using gProfiler. Pathways are ranked by adjusted p-value (Padj) after multiple-testing correction and include functional categories from Gene Ontology: Molecular Function (GO:MF) and Biological Process (GO:BP).

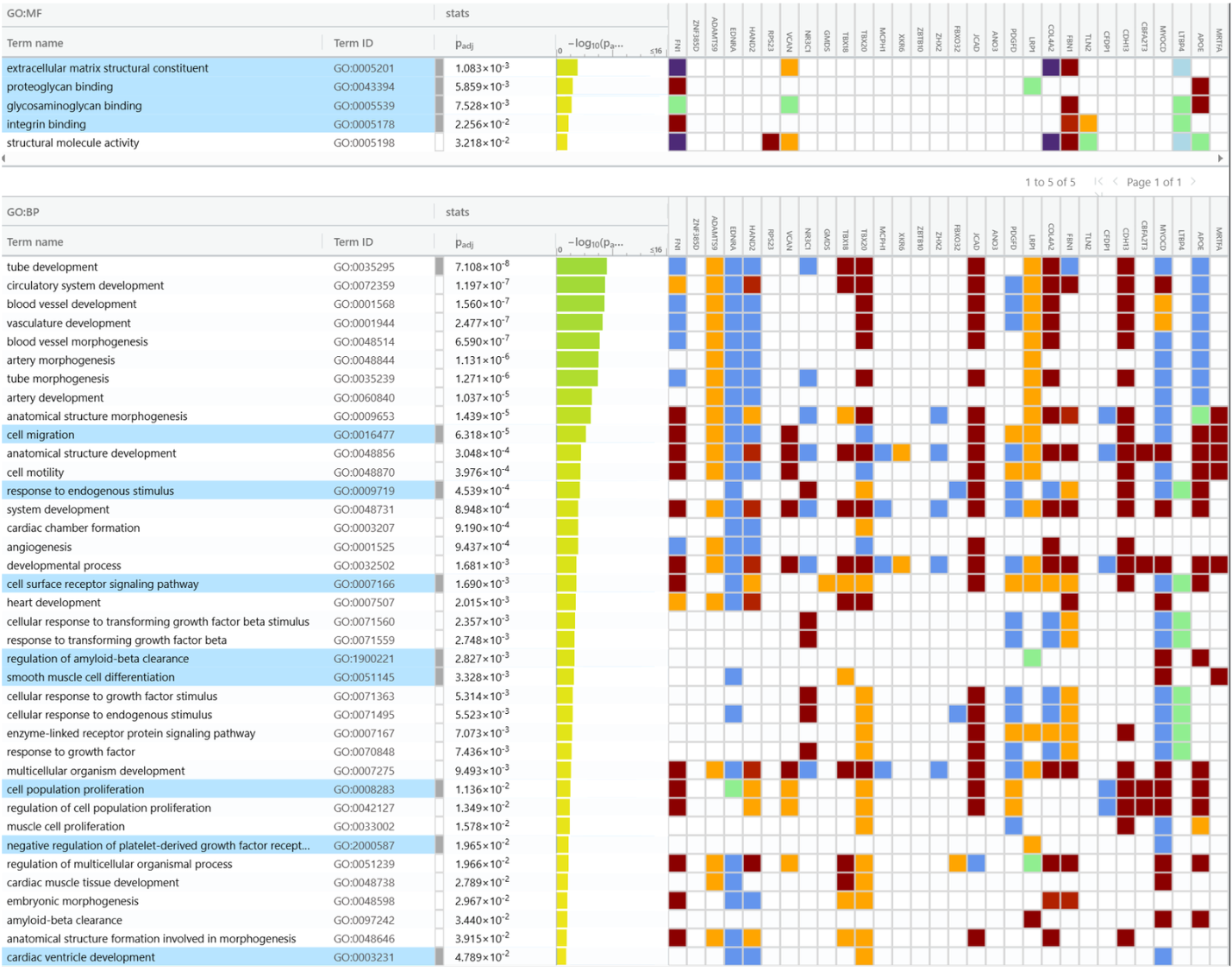
